# EIT-based non-invasive measurements correlate with pulmonary arterial pressure in patients

**DOI:** 10.64898/2026.10.30.26366927

**Authors:** Jonathan Boudreau, Frédérick Drouin, Mathieu Rivet, Carlos Fallaque, Jonathan Toma, Jean-Marc Lina, Andy Adler

## Abstract

**Objective:** To investigate associations between electrocardiogram (ECG)-referenced timing markers derived from electrical impedance tomography (EIT) and invasively measured mean pulmonary artery pressure (mPAP) in patients with cardiovascular disease.

**Approach:** EIT, ECG and pulmonary artery pressure were recorded in seven patients during a protocol combining apnoea and phenylephrine administration. Four waveform landmarks were considered: the maximum, the maxima of the first and second derivatives, and the tangent intersection. Their delays from ECG R waves defined eight markers across the heart and lung regions. Within-patient Pearson and Spearman correlations were calculated against unsmoothed invasive mPAP.

**Main results:** Across 56 patient–marker combinations, the mean absolute Spearman coefficient was 0.74. Absolute coefficients reached 0.70 in 40 combinations and 0.80 in 27. Each patient had at least one marker with an absolute coefficient above 0.79. The lung second-derivative timing marker exceeded 0.90 in four patients, more often than any other marker, but showed weaker associations in the remaining three. Associations were positive in 25 combinations and negative in 31. Their strength and direction varied across patients and landmarks.

**Significance:** These exploratory findings support studying regional EIT timing as a non-invasive correlate of pulmonary haemodynamics. Validation in independent recordings is required before using these markers to estimate pressure. EIT timing markers are associated with PAP changes in patients; however, there is significant variation between patients.

## 1 Introduction

Pulmonary artery pressure (PAP) is an important descriptor of cardiac and pulmonary haemodynamic status. The PAP waveform reflects right-heart loading, and changes in PAP can provide an early indication of worsening congestive heart failure (CHF). In CHF associated with left heart disease, elevated left-sided filling pressures can be transmitted to the pulmonary circulation, increasing PAP (Humbert et al., 2022). Zile et al. (2008) observed pressure elevations before clinical decompensation, and Zalawadiya et al. (2026) found that mean PAP rose weeks before heart failure events while weight, systolic blood pressure and heart rate did not show consistent changes.

The gold standard for PAP measurement uses a Swan–Ganz catheter advanced through the venous system into the pulmonary artery (Swan et al., 1970; Humbert et al., 2022). Catheterization requires invasive vascular access and carries procedural risks (Hoeper et al., 2006). Ultrasound-based estimates of systolic PAP use tricuspid regurgitation velocity and estimated right atrial pressure (Humbert et al., 2022; Fisher et al., 2009). These assessments require trained operators and provide intermittent measurements (Proença et al., 2020). Implantable pressure sensors, such as CardioMEMS, enable repeated monitoring outside the catheterization laboratory. Although they reduce the need for repeated catheterization, implantation still requires a procedure and carries a risk of complications (Gibson et al., 2023).

There is therefore a need for continuous, non-invasive approaches to PAP monitoring. Electrical impedance tomography (EIT) uses body-surface electrodes to image dynamical changes in thoracic impedance (Frerichs et al., 2017). EIT-based PAP estimation has been proposed using changes in the timing of cardiac-related pulmonary signals (Proença et al., 2016, 2020). The physiological rationale involves pulse-wave velocity (PWV), the speed at which the pressure wave travels through the pulmonary arteries. Higher PAP is associated with reduced arterial distensibility and shorter pulmonary transit times (Lau et al., 2012; Mueller-Graf et al., 2021).

Studies in healthy volunteers and experimental animals support this approach. An initial human study examined pulmonary EIT timing during induced hypoxia (Proença et al., 2016). A subsequent study analysed 24 healthy participants using two-minute ECG-gated ensembles and calibrated timing against echocardiographic systolic pulmonary artery pressure (Proença et al., 2020). Direct pulmonary transit measurements in a six-pig experiment also showed an inverse relationship with pulmonary artery pressure, using invasive sensors rather than ECG-referenced EIT (Mueller-Graf et al., 2021).

EIT-based PAP measurement is difficult because cardiac-related signals are small and cardiac and pulmonary contributions mix within the EIT image (Grant et al., 2011; Frerichs et al., 2017). Their separation can depend on anatomy and lung ventilation. The signal still mixes blood-volume changes, cardiac motion and reconstruction effects; neither a regional impedance change nor its timing directly measures pressure. Clinical EIT studies have investigated pulmonary pulsatility amplitude in patients with pulmonary arterial hypertension (Hovnanian et al., 2021). However, the feasibility and reliability of regional ECG-referenced timing markers remain incompletely characterized in surgical patients with cardiovascular disease.

To our knowledge, this is the first study to report associations between regional ECG-referenced EIT timing markers and invasively measured mean pulmonary artery pressure (mPAP) in surgical patients with cardiovascular disease. Our goal is to investigate the feasibility and reliability of EIT timing markers for tracking PAP changes in this clinical population. EIT signals with synchronized ECG were recorded in seven patients whose clinical care already included planned surgery with Swan–Ganz catheterization. Phenylephrine was administered during apnoea to induce haemodynamic changes. Four waveform landmarks were calculated in each of the heart and lung regions, providing eight ECG-referenced timing markers. We assess detection quality, variability across patients and marker-specific associations with invasive mPAP.

## 2 Methods

### 2.1 Patients and data collection

Data were collected from seven patients undergoing surgery at Southlake Regional Health Centre, Ontario, Canada, with approval from the Southlake Regional Health Centre Research Ethics Board (protocol S-018-2122). All patients had underlying cardiac disease and were already scheduled for surgery with Swan–Ganz catheterization as part of their clinical care. The surgery and catheter placement had been planned by the treating clinical team independently of the study. During the pre-surgical preparation, in addition to the ECG, patients were fitted with an EIT belt for data collection using a PulmoVista 500 system (Dräger Medical, Lübeck, Germany).

The dataset comprised EIT recordings, R-wave timing derived from ECG lead I, videos of the clinical monitor and manually transcribed mPAP values. These recordings provided the spatial impedance signals, cardiac timing reference and invasive pressure reference required for the analysis.

Table 1 summarizes the demographic and clinical characteristics of the seven patients included in the study. Left ventricular measurements were obtained by transthoracic echocardiography, and PR and QRS intervals were obtained from surface ECG. The recorded cardiovascular conditions included coronary artery disease, valvular disease, an ascending aortic aneurysm and ventricular dysfunction.

**Table 1.** Demographic and clinical characteristics of the seven patients.

| Characteristic | Mean ± SD or count | Range | *n* |
|---|---|---|---|
| Age (years) | 66.7 ± 9.4 | 52–79 | 7 |
| Sex (male/female) | 6/1 | — | 7 |
| LVEF (%) | 50.8 ± 14.6 | 25–67 | 6 |
| LVEDD (mm) | 47.2 ± 3.6 | 42–52 | 6 |
| PR interval (ms) | 183.7 ± 37.3 | 148–250 | 7 |
| QRS duration (ms) | 89.4 ± 14.2 | 70–112 | 7 |

SD denotes the sample standard deviation; *n* denotes the number of patients with an available value. LVEF: left ventricular ejection fraction; LVEDD: left ventricular end-diastolic dimension. LVEF and LVEDD were not reported for one patient. Sex is reported as counts; — denotes not applicable.

Measurements were acquired with patients in the supine position. The original protocol included controlled apnoea and intravenous phenylephrine administration to introduce haemodynamic variation during acquisition. Apnoea reduced respiratory impedance fluctuations, allowing the cardiac-related component to be examined. The present analysis selected cycles with R-wave anchors inside the documented apnoea intervals and focused on regional waveform landmark extraction and its association with mPAP. The final selected cycle could extend beyond the annotated end of apnoea. Associations were examined within each recording to describe variability across patients. Figure 1 illustrates the sequence of apnoea, phenylephrine injection and pulmonary artery pressure variation.

**Figure 1.**
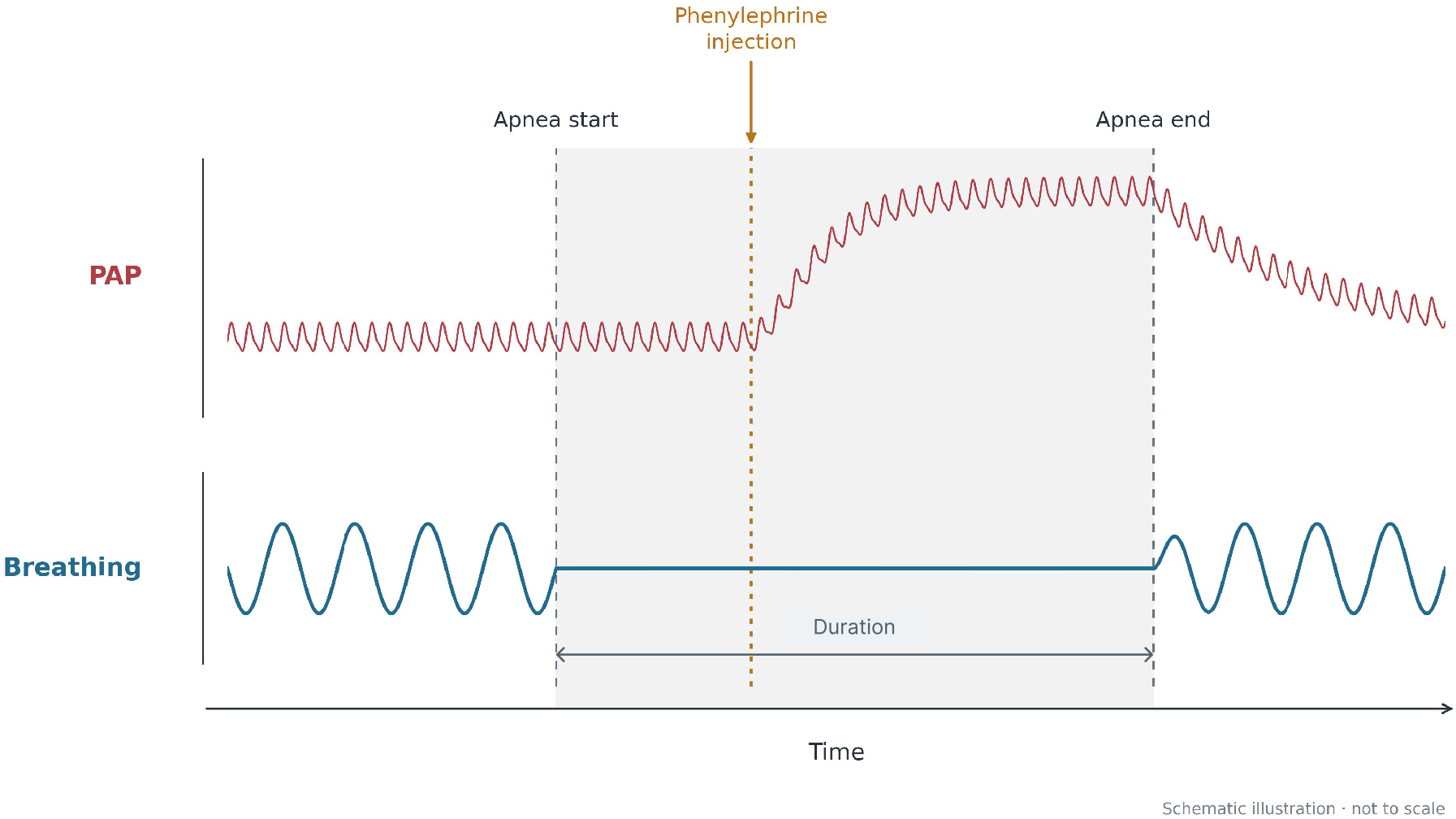
Schematic timeline of the experimental protocol. Grey dashed lines delimit apnoea (shaded interval), and the orange marker denotes phenylephrine injection. The upper curve illustrates a pulmonary artery pressure (PAP) increase following injection and a later decrease; the lower curve represents breathing, interrupted during apnoea. Amplitudes and durations are illustrative and not to scale.

Table 2 summarizes the main cardiovascular conditions represented in the cohort.

**Table 2.** Main cardiovascular conditions of the seven patients.

| Patient | Main cardiovascular conditions |
|---|---|
| 1 | Coronary artery disease. |
| 2 | Coronary artery disease; aortic stenosis. |
| 3 | Coronary artery disease; ventricular dysfunction. |
| 4 | Prosthetic aortic valve stenosis. |
| 5 | Ascending aortic aneurysm; coronary artery disease. |
| 6 | Aortic stenosis. |
| 7 | Coronary artery disease. |

### 2.2 Acquisition, pressure reference and synchronization

EIT data were acquired using a PulmoVista 500 system (Dräger Medical, Lübeck, Germany), with a belt of 16 electrodes placed around the thorax at the mid-thoracic level. The EIT acquisition rate was 50 frames s^*−*1^, corresponding to a nominal sampling interval of 20 ms. ECG was acquired at 250 Hz, corresponding to a sampling interval of 4 ms. ECG R-wave peaks provided the temporal anchors for cardiac-cycle segmentation and waveform timing.

Pulmonary artery pressure was measured invasively using a Swan–Ganz catheter. The reference available for this analysis was the monitor-displayed mPAP, manually transcribed from the monitor video at one-second timestamps. It was therefore a scalar pressure series sampled at 1 Hz, rather than a digitized pulmonary artery pressure waveform. Figure 2 summarizes the acquisition setup and the temporal resolution of the available signals.

**Figure 2.**
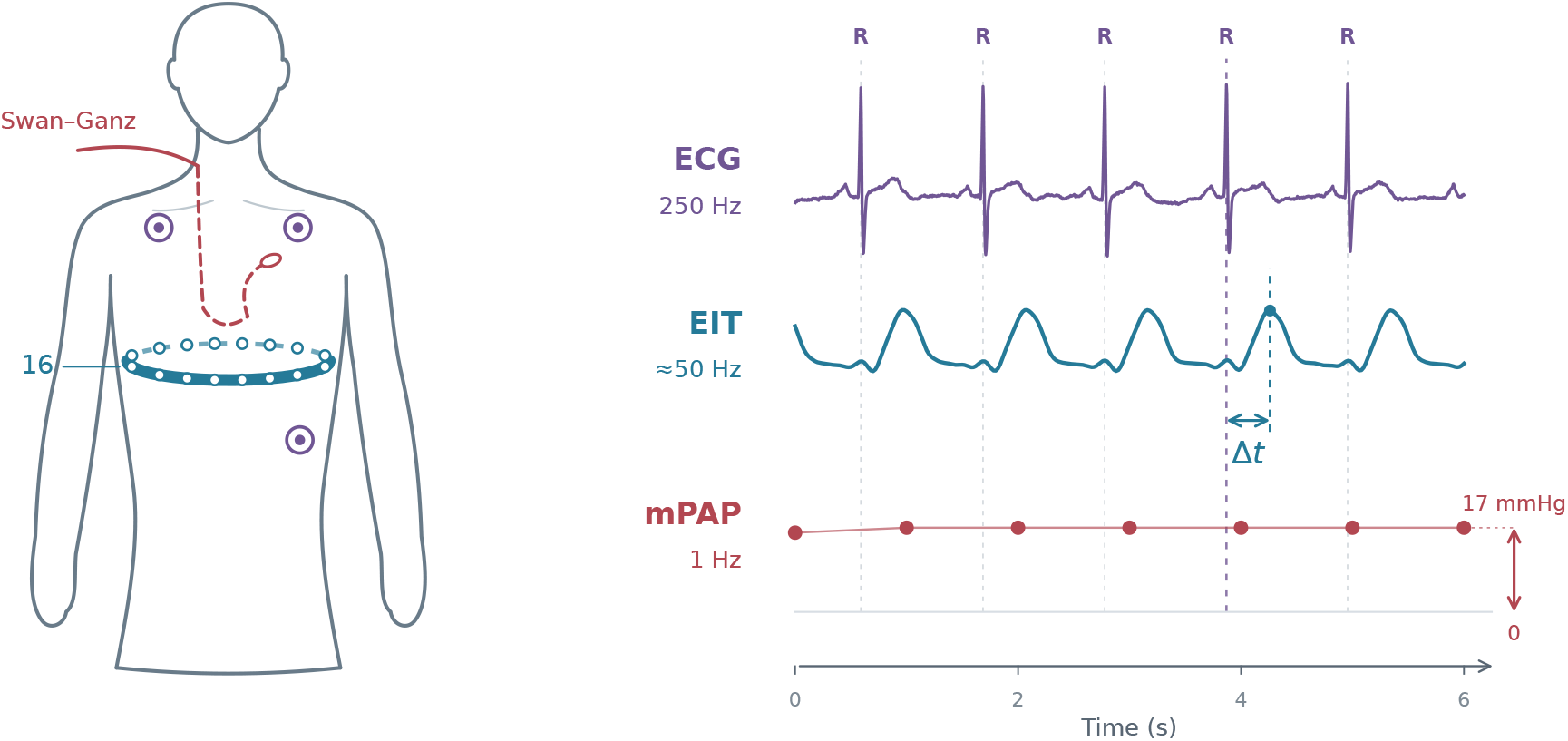
Acquisition setup and representative archived signals. The body, electrode positions and Swan–Ganz catheter path are schematic. The six-second apnoea excerpt shows native ECG (250 Hz), the spatial mean of cardiac-band EIT signals (0.7–7 Hz; 50 frames s^*−*1^) in a fixed functional lung mask, and monitor-displayed mPAP transcribed at 1 Hz. ECG and EIT amplitudes are independently scaled for display. The blue arrow illustrates the delay from a native ECG R peak to a regional-mean EIT maximum; this differs from the pixel-wise median CP-WD endpoint. The red arrow indicates the scalar mPAP level of 17 mmHg above zero, not pulse pressure. Pressure points are native observations, with straight segments as visual guides.

The EIT, ECG and pressure signals were already synchronized.

### 2.3 Image reconstruction and regional signal definition

EIT images were reconstructed using the GREIT algorithm implemented in EIDORS (Adler et al., 2009; Adler and Lionheart, 2006). The reconstruction used a thoracic finite-element model configured for the 16-electrode acquisition geometry. Image values represented changes relative to an electrical reference, and each reconstructed pixel provided a time series at the EIT frame rate. The cardiac-related component was isolated by filtering the pixel time series in the 0.7–7 Hz band. Figure 3 summarizes the reconstruction and regional-signal workflow.

**Figure 3.**
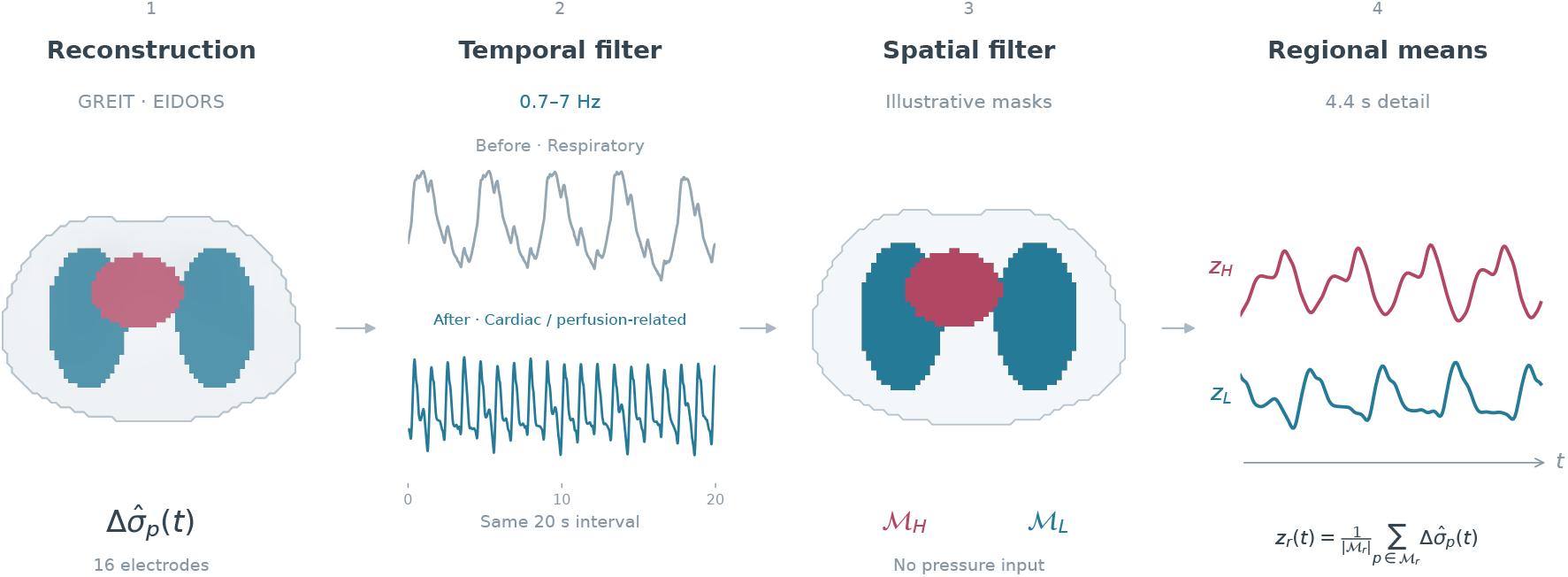
Reconstruction and regional-signal workflow: (1) GREIT reconstruction in EIDORS for the 16-electrode geometry; (2) cardiac-band filtering (0.7–7 Hz); (3) generic functional heart and lung masks illustrating spatial selection; (4) regional mean waveforms for visualization. Panel 2 shows the same 20-s lung-region interval before and after filtering; panel 4 shows a 4.4-s detail from that interval. The displayed *z*_*H*_ is the filtered heart regional mean before the sign reversal used for timing analysis.

Heart (H) and lung (L) regions of interest (ROIs) were determined algorithmically from the reconstructed EIT signals. The lung masks were constructed from the amplitude of respiratory-related impedance variations in the 0.1–0.5 Hz band, measured before and after apnoea. The heart mask was then determined outside the lung regions from the periodic activity of signals aligned to the ECG R waves. Selected pixels were grouped into connected regions to form patient-specific binary masks, denoted ℳ_*H*_ and ℳ_*L*_. These masks were held fixed throughout the analysed interval, so that successive cardiac cycles were evaluated using the same spatial support. Pulmonary artery pressure was not used in this selection.

For a region *r ∈ {H, L}*, the spatially averaged waveform was defined as

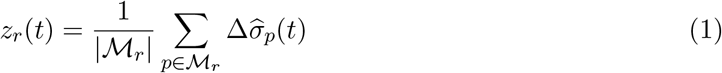

where 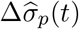 denotes the reconstructed conductivity change at pixel *p* and |*ℳ*_*r*_| is the number of pixels in the mask. Regional averages summarized the waveform morphology for visualization. Timing-marker extraction and the aggregation of pixel delays are described separately in the next subsection.

ECG R-wave anchors provided a common cardiac-phase reference for comparing the regional waveforms. The archived recording-level polarity convention was applied to both regions, followed by a fixed sign reversal of the heart signal before timing analysis. The heart and lung masks identified regions according to the activity observed in the EIT signals. They therefore did not necessarily represent the exact anatomical outlines of the heart and lungs.

### 2.4 Definition and extraction of the eight CP-WD markers

Cardiopulmonary waveform delay (CP-WD) was defined as the interval from an ECG R-wave anchor to a specified landmark of the reconstructed EIT waveform. For cardiac cycle *k*, region *r* and pixel *p*, the delay associated with landmark *m* was

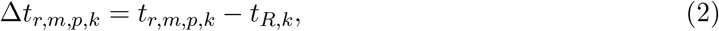

where *t*_*R,k*_ is the ECG R-wave time and *t*_*r,m,p,k*_ is the landmark time. Delays were expressed in milliseconds, with a positive value indicating a landmark after the R wave. The R wave provides an electrical timing anchor; the measured interval can include electromechanical timing, propagation and waveform-formation effects. Figure 4 illustrates the four landmark definitions and their common ECG timing anchor.

**Figure 4.**
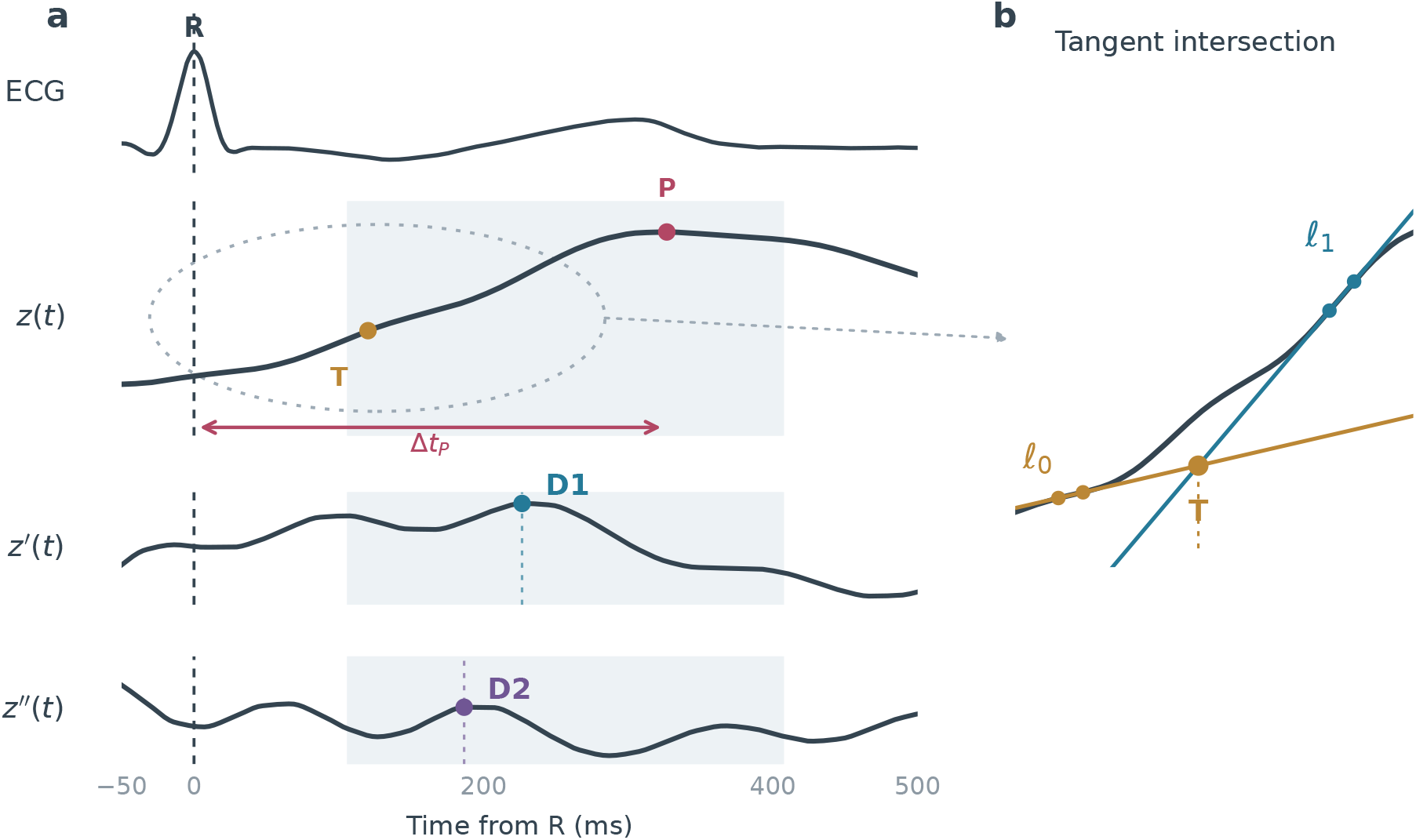
Definition of the four CP-WD landmarks using recorded ECG and cardiac-band EIT signals from one observed cycle. (a) The native ECG R-wave maximum defines time zero for this illustration; the ECG T wave is labelled separately from the EIT tangent marker T. P, D1 and D2 mark native-sample maxima of *z*(*t*) and its first and second derivatives within the shaded example interval. Numerical extraction searches the full observed R–R interval and refines extrema as described in the text. The arrow shows the R-to-P delay; the dashed oval identifies the segment enlarged in panel (b). (b) T illustrates the intersection of two forward secants with a common 20 ms interval; the detector uses local linear fits. A common time translation aligns all signals to the native R maximum; archived analysis anchors and EIT landmark extraction remain unchanged. Numerical precision does not establish sub-frame measurement accuracy.

Landmarks were detected separately in each pixel of the fixed heart and lung masks. Let *Z*_*r,p,k*_(*t*) denote the cardiac-related pixel impedance signal after cardiac-band filtering and the polarity convention described in section 2.3. The waveform maximum (P), maximum first derivative (D1) and maximum second derivative (D2) were defined as

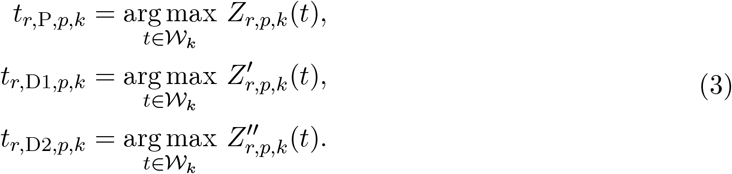

Here, *W*_*k*_ = [*t*_*R,k*_, *t*_*R,k*+1_) is the observed interval between successive R-wave anchors. Intervals without a following R anchor, at least five EIT samples or continuous acquisition support were unavailable. The numerical temporal derivatives *Z*^*′*^ and *Z*^*′′*^, calculated with second-order gradient formulae. Extrema were found with a local three-sample parabola fit when its vertex lay between the neighbouring samples; otherwise the sampled extremum time was retained. The same definitions were applied to both regions.

The tangent-intersection marker (T) used two local least-squares lines: one fitted around the minimum preceding the upstroke and the other around the maximum first derivative occurring no later than P. The fits used up to three and five adjacent EIT samples, respectively, with the upstroke fit ending no later than the sampled peak. Writing the two lines as *l*_0_(*t*) = *a*_0_*t* + *b*_0_ and *l*_1_(*t*) = *a*_1_*t* + *b*_1_, their intersection time was

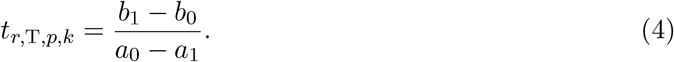

A finite intersection required non-parallel lines, and the tangent marker had to lie in the observed cardiac cycle, at or before the sampled waveform maximum. The line intersection defines a geometric timing estimate; its numerical precision does not establish sub-frame measurement accuracy.

After landmark detection, the pixel delays were aggregated within each region using their median:

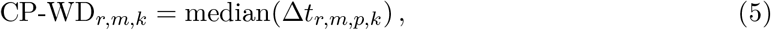

A regional value was not used when calculated pixel delay was outside of bounds (0,500 ms). The two regions *r ∈ {H, L}* and four landmarks *m ∈ {*P, D1, D2, T*}* therefore yielded eight CP-WD markers. No additional temporal filter was applied to the regional delay series or the pressure reference. Figure 5 summarizes the pixel-wise delay aggregation for the two regions and four landmarks.

**Figure 5.**
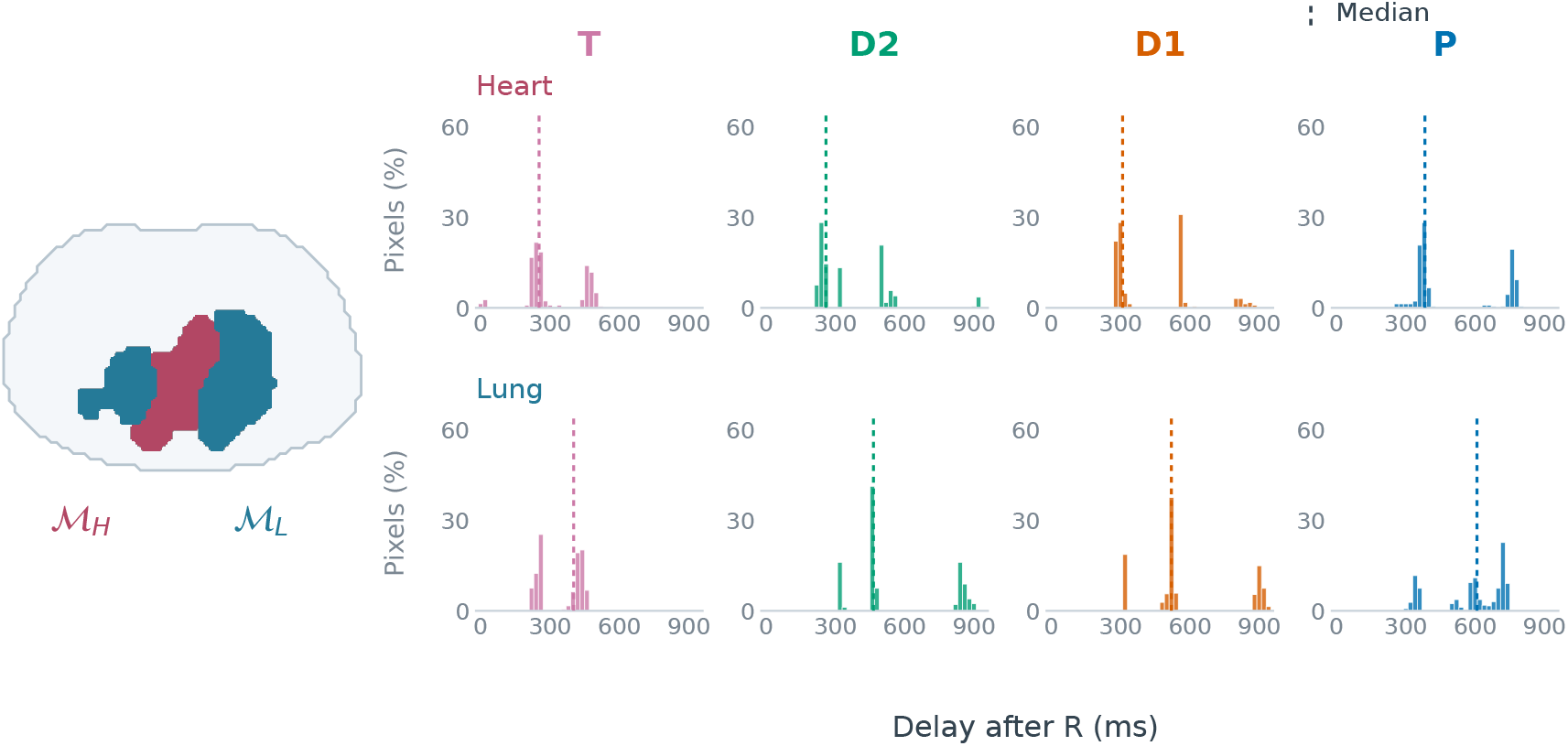
Recorded functional masks and pixel-delay distributions from one observed cycle in one patient. All 226 heart-region and 428 lung-region native pixels are included for each marker. Finite delays were copied unchanged from the frozen full-cycle source, without repeating landmark extraction. Bars show percentages of finite pixels in 20 ms bins; dashed lines indicate unbinned medians. Columns are ordered T, D2, D1 and P.

### 2.5 Correlation analysis

For each patient, we compare each of the eight CP-WD markers with mPAP. Spearman’s coefficient uses their ranks to describe an increasing or decreasing trend, which need not follow a straight line.

A positive coefficient indicates changes in the same direction; a negative coefficient indicates opposite changes. The closer the coefficient is to +1 or *−* 1, the stronger the measured association. We report the signed coefficients and the number *n* of available pairs. These correlations do not measure the accuracy of pressure estimates.

## 3 Results

All eight CP-WD markers were available in the seven patients, yielding 56 patient–marker combinations. For each combination, Pearson and Spearman coefficients were calculated against unsmoothed invasive mPAP.

Figure 6 compares all four archived CP-WD components in the lung and heart regions of patient 1 with the pressure reference over the complete annotated apnoea interval. The signed coefficients and observation counts use the same stored series and pairing as the correlation figures.

**Figure 6.**
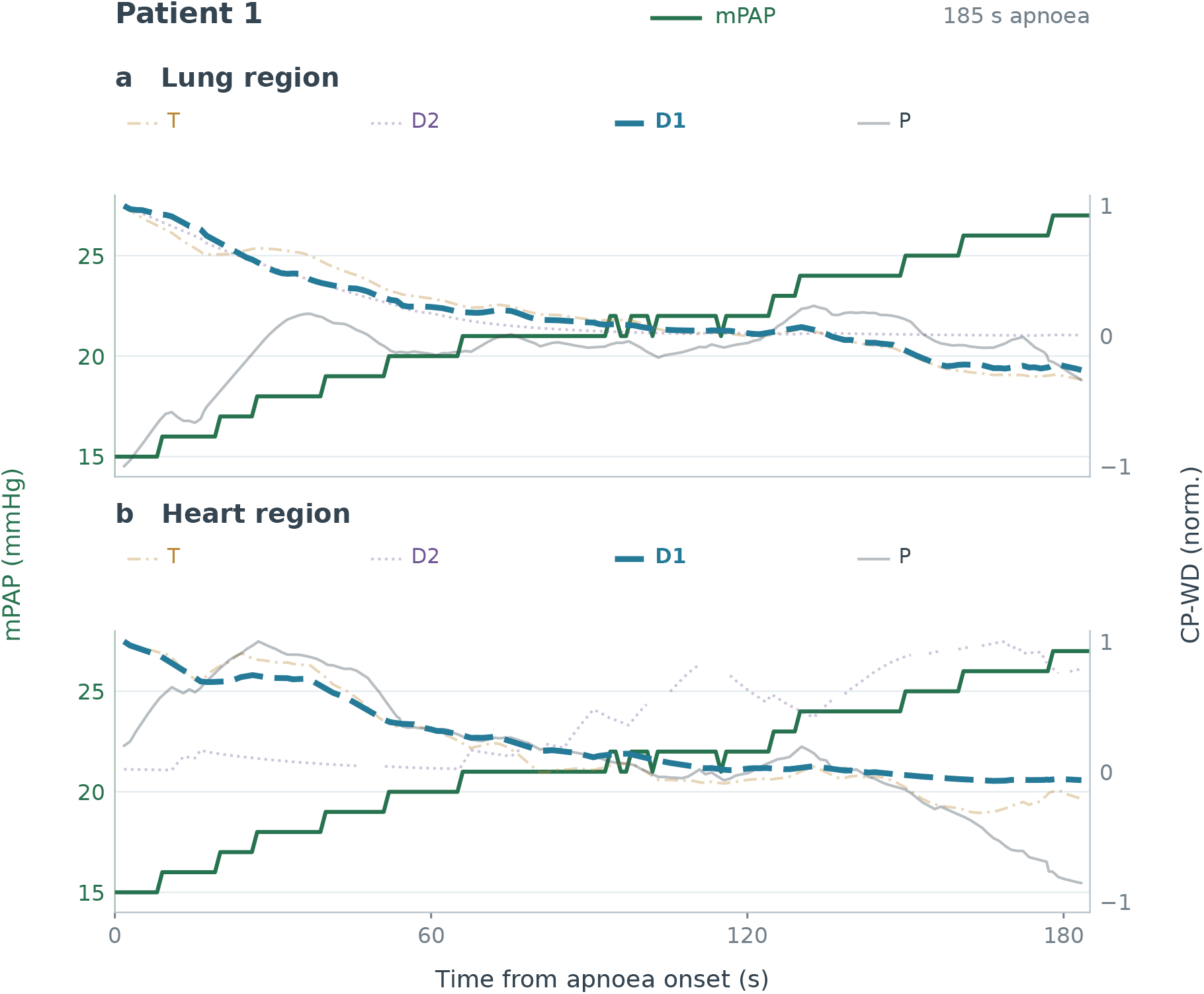
Patient-1 CP-WD series and unsmoothed invasive mPAP throughout the 185-s apnoea interval. Panels show (a) lung and (b) heart T, D2, D1 and P. D1 is emphasized in each region because it has the largest absolute regional Spearman coefficient; all markers remain visible. Pressure is displayed at native 1-Hz timestamps in mmHg. Each CP-WD trace is divided by its positive maximum absolute finite paired value, preserving zero, signs and original pair masks. Gaps in the CP-WD traces correspond to points with insufficient signal quality.

Table 3 reports all signed Spearman coefficients. The mean absolute Spearman coefficients were 0.74, respectively. Both magnitudes reached 0.70 in 38 of 56 combinations and 0.80 in 22 of 56. These descriptive counts include every patient and marker.

**Table 3.** Signed Spearman associations between the eight CP-WD markers and unsmoothed invasive mPAP for all seven patients. Within each region, columns are ordered T, D2, D1 and P. All 56 signed coefficients are printed to three decimal places. Within each patient, the largest absolute coefficient among the eight measures is bold, using the unrounded coefficients to select the maximum.

|  | **Lung** |  |  |  | **Heart** |  |  |  |
|---|---|---|---|---|---|---|---|---|
|  | T | D2 | D1 | P | T | D2 | D1 | P |
| Patient 1 | -0.983 | -0.983 | **-0.984** | +0.305 | -0.946 | +0.891 | -0.980 | -0.923 |
| Patient 2 | -0.885 | -0.514 | -0.843 | +0.783 | -0.848 | +0.345 | **-0.920** | +0.526 |
| Patient 3 | +0.526 | +0.698 | +0.476 | -0.738 | +0.770 | **+0.796** | +0.770 | +0.744 |
| Patient 4 | -0.710 | +0.548 | -0.592 | **-0.917** | +0.877 | +0.824 | +0.896 | -0.092 |
| Patient 5 | -0.954 | **-0.959** | -0.951 | +0.664 | -0.851 | +0.858 | -0.732 | +0.943 |
| Patient 6 | -0.882 | **+0.908** | -0.798 | +0.787 | -0.191 | +0.226 | -0.601 | +0.808 |
| Patient 7 | -0.777 | +0.952 | -0.918 | -0.231 | **-0.954** | +0.749 | -0.754 | -0.427 |

Bold: largest \|ρ\| within each patient

Spearman signs agreed in every combination: 25 associations were positive and 31 negative. Direction therefore varied across patients and landmarks. In patient 1, lung D1 yielded *ρ* = *−* 0.98, and lung T yielded *ρ* = *−* 0.98. In patient 4, lung P yielded *ρ* = *−* 0.92. Strong magnitudes were not uniform across markers or patients, and their signed grand means were close to zero because opposite directions partly cancelled. These results describe association within the observed episodes, rather than agreement of estimated and measured pressure.

Figure 7 shows the paired observations for patients 1, 2 and 6, selected by their highest, median and lowest mean absolute Spearman coefficient across the eight markers. The scatterplots illustrate differences in direction and shape between markers within these recordings; the selection does not summarize the population.

**Figure 7.**
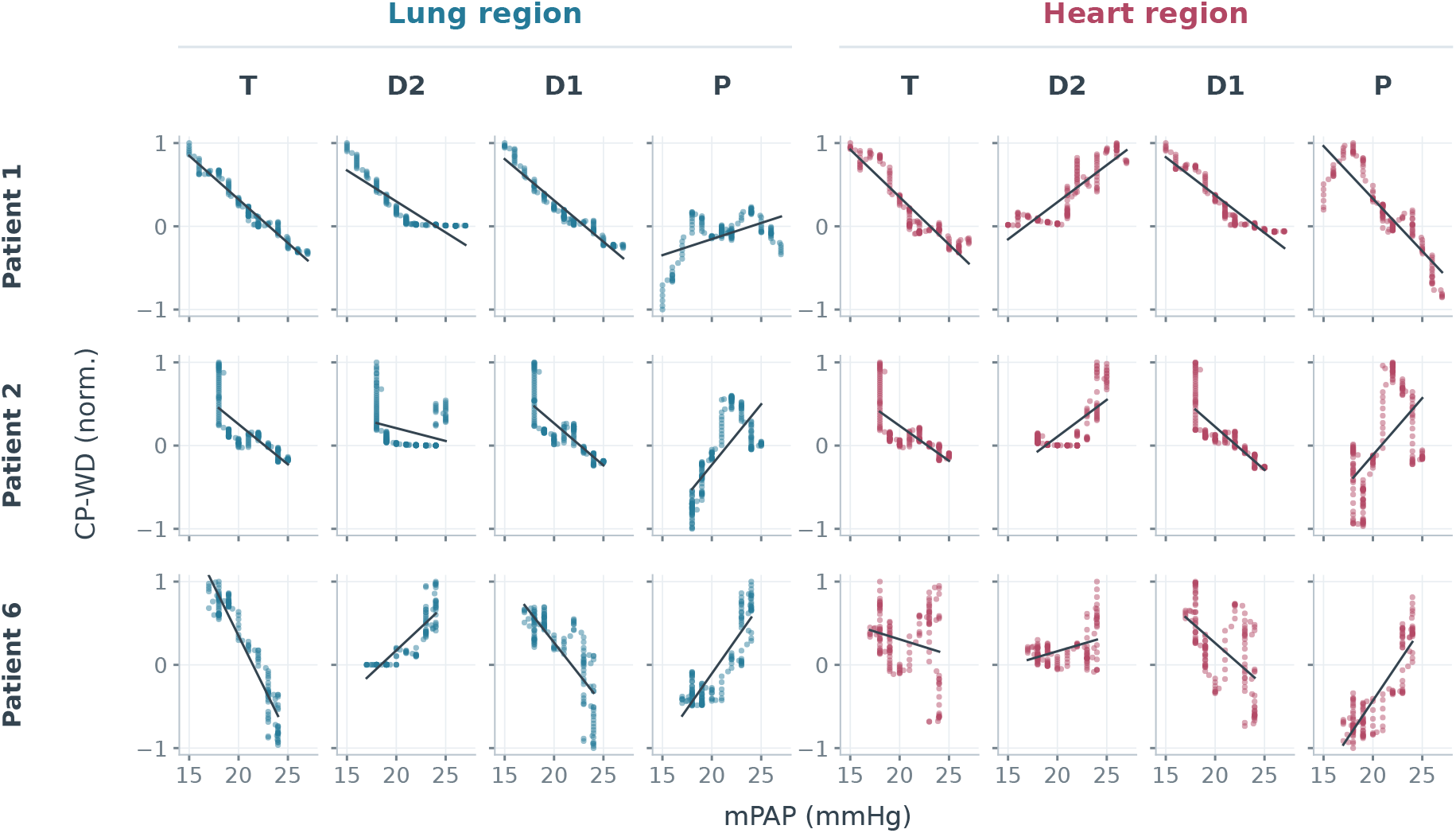
Within-patient associations between unsmoothed invasive mPAP and CP-WD for patients 1, 2 and 6. Columns show lung T, D2, D1 and P, followed by heart T, D2, D1 and P. Every point is an original finite statistical pair, using stored pressure interpolation at CP-WD timestamps. Each processed CP-WD series is divided by its own positive maximum absolute paired value, preserving signs and zero. Black lines are descriptive ordinary-least-squares fits of displayed CP-WD on mPAP.

## 4 Discussion

### 4.1 Main findings

Our goal was to investigate whether EIT timing markers could track changes in pulmonary artery pressure in patients with cardiovascular disease. We extracted eight CP-WD markers in seven patients during apnoea with phenylephrine administration, then compared their changes with invasive mPAP.

The mean absolute Spearman coefficient, *ρ* = 0.74 across the 56 patient–marker combinations. We observed strong associations for several markers. Across the seven patients and both regions, D1 had the highest mean absolute Spearman coefficient 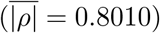, followed by T (0.7966), D2 (0.7322) and P (0.6350). The lung D2 marker had an absolute coefficient above 0.90 in four patients. However, the direction and strength of these associations changed between patients. No marker gave the best results in every case.

To our knowledge, this is the first study to report these timing-marker associations with invasive mPAP in patients. Earlier work by Proença et al. (2020) examined healthy volunteers during controlled hypoxia. These first clinical results show that associations can be obtained in patients with cardiovascular disease, while also revealing substantial variability between patients.

### 4.2 Physiological significance of the delays

To understand these associations, we need to consider the phenomena that contribute to the measured delay. CP-WD starts at the ECG R wave. It therefore includes the time between electrical activation and blood ejection, as well as the processes of propagation and EIT waveform formation. The importance of pre-ejection timing has been shown in the systemic circulation, but its contribution to the pulmonary markers used here was not measured (Payne et al., 2006).

The EIT signal can also contain several contributions, including blood volume changes and the mechanical effects of the heart on surrounding tissue. These contributions may differ between the heart and lung regions (Grant et al., 2011).

A marker can therefore respond to haemodynamic changes without all its variations being caused by PAP. Our results show associations with pressure, but do not separate the different physiological contributions. To improve marker specificity, we will need to better identify the signal components related to the pulmonary circulation.

### 4.3 Example of waveform morphology

We illustrate the results in a patient in (figures 6 and 7). In the lung region, T, D1 and D2 generally move closer to the R wave as mPAP increases. Peak P does not follow the same decrease.

Some associations are positive, and others negative. One possible explanation would be a waveform whose upstroke begins earlier while its peak changes little in time. Landmarks on the upstroke could then move earlier by more than the maximum. The time between the start of the upstroke and the peak would become longer. This behaviour could explain why the markers show different trajectories.

In the heart region, T and D1 generally become earlier, whereas D2 becomes later. This could correspond to a change in waveform slope and curvature.The trajectories shown illustrate the differences between landmarks, but are not sufficient to confirm the proposed changes in morphology.

### 4.4 Choice of the spatial mask

The mask determines which pixels are used to calculate regional CP-WD. Its choice can therefore change the result, and a careful selection of the mask was necessary by choosing the phase, frequency and amplitude thresholds. The limited resolution of EIT also complicates the separation of signals from the heart and lungs. A functional mask selects activity observed in the image; its boundaries do not necessarily correspond to anatomical boundaries (Grant et al., 2011).

In our study, masks were specific to each patient and remained fixed throughout the analysed interval. Their definition did not use PAP. However, some pixels may provide a landmark in one cycle and not in the next. Because the regional delay is calculated as the median of the available delays, these changes can affect its value.

### 4.5 Limitations and next steps

This study includes seven patients with different cardiovascular diseases. Anatomical differences and comorbidities could contribute to the variability in the results. However, the number of patients remains too small to determine the effect of each disease or to generalise the observed behaviour. The mPAP reference at 1 Hz also limits the analysis of timing relationships at each heartbeat.

### 4.6 Conclusion

Regional EIT–ECG timing markers showed substantial associations with unsmoothed invasive mPAP in seven recordings, with patient- and landmark-dependent directions and magnitudes. This study shows that EIT timing markers are associated with PAP changes in patients. The results are encouraging, but differences between patients remain substantial. Further work will need to better explain these differences and determine which landmarks and regions provide the most stable measurements.

## Ethical statement

The study was approved by the Southlake Regional Health Centre Research Ethics Board, Ontario, Canada (protocol S-018-2122; approval dated 27 October 2021; protocol version 0.2). The research was conducted in accordance with the principles of the Declaration of Helsinki and applicable local statutory requirements. All participants provided written informed consent to participate in the study.

## Data availability statement

The clinical data and analysis code supporting this study are not publicly available.

